# Analysis of Integrated Management of Acute Malnutrition (IMAM) dataset 2013 to 2018: A secondary dataset analysis

**DOI:** 10.1101/2023.02.09.23285728

**Authors:** V. Kamazizwa, T.K Nyadzayo, G. Shambira, N.T Gombe, T. Juru, M. Tshimanga

## Abstract

**Background:** Malnutrition remains the most common cause of morbidity and mortality in children under the age of 5 years. In Zimbabwe nearly 1 in every 3 children is malnourished. Child nutrition in the country is affected by inadequate knowledge among mothers and caregivers on appropriate diets especially in the first 1000 days of life. Weak coordination and inadequate resources for high impact nutrition interventions also affect nutrition in children.

**Methods:** A descriptive cross sectional study was conducted on records from January 2013 to December 2018. All malnourished patients admitted into Community based management of acute malnutrition (CMAM) at health facilities in, with a diagnosis of Severe Acute Malnutrition (SAM) from January 2013 December 2018 as reported in the DHIS2 were included in the study. We retrieved the dataset from the DHIS2 to the excel sheet and analysed the data to come up with graphs demonstrating trends in management of acute malnutrition.

**Results:** The 6-59 months age-group constitute the group with the highest cases of admission into the CMAM program. The 0-5 months age-group showed the least number of cases admitted and had the lowest HIV positivity over the period under review. The highest treatment outcome for HIV positive patients was death rate. Highest number of cases of malnutrition were admitted in 2016.The District failed to meet sphere standard default and cure rate throughout the period under review.

**Conclusion:** The treatment outcomes of this study showed that HIV negative had better treatment outcomes than HIV positive. The age group 6 to 59 months had the highest number of cases admitted into CMAM program.The implementation of an effective public health approach for addressing malnutrition require a holistic approach that incorporates preventive measures such as poverty reduction, hygiene promotion, clean water, prevention of prenatal macronutrient and micronutrient deficiencies, and the promotion of age appropriate foods and feeding practices for infants and young children.

## Introduction

Malnutrition refers to deficiencies, excesses or imbalances in a person’s intake of energy and nutrients(1).Malnutrition remains one of the most common causes of morbidity and mortality among children throughout the world. Globally it is estimated that 35% of all child mortality is attributable to maternal and child under nutrition(2).

Acute malnutrition is caused by a decrease in food consumption and illness resulting in bilateral pitting oedema or sudden weight loss. It is defined by the presence of bilateral pitting oedema or wasting(3).Anorexia, or poor appetite, and medical complications are clinical signs indicating or aggravating the severity of acute malnutrition(4).

There are two forms of acute malnutrition which are severe acute malnutrition (SAM) and moderate acute malnutrition (MAM). SAM is defined by the presence of bilateral pitting oedema or severe wasting, and other clinical signs such as poor appetite. A child with SAM is highly vulnerable and has a high risk of death. MAM is defined by moderate wasting(4).

The clinical manifestations of SAM are marasmus, kwashiorkor and marasmic kwashiorkor. Marasmus is characterised by severe wasting of fat and muscle, which the body breaks down for energy, leaving skin and bones. Kwashiorkor is characterised essentially by bilateral pitting oedema which usually start in the feet and legs, accompanied by a skin rash and/or changes in hair colour (greyish or reddish). Marasmic kwashiorkor is characterized by a combination of severe wasting and bilateral pitting oedema(4).

Globally it is estimated that 35% of all child mortality is attributable to maternal and child undernutrition. Severe acute malnutrition remains a major killer of children under five years old. It contributes to 1.7 million child deaths per year in sub Saharan Africa. The risk of death in children with severe acute malnutrition (SAM) is 10 times greater than the risk of death in their well-nourished counterparts and the risk of death in children with moderate acute malnutrition is more than 2 times greater than in their well-nourished counterparts(2).

According to the national nutrition survey (unpublished) of 2018 the prevalence of global acute malnutrition (GAM) is 0.2 %. The prevalence of MAM (<-2 z score and >= -3 z score, no oedema) is 2.3 % and the prevalence of severe acute malnutrition (<-3 z score and / oedema) is 0.2%. The global threshold for acute malnutrition is >15% GAM or 10 to 14 % with aggravating factors according to WHO standards.

In order to identify children with malnutrition, the Ministry of Health and Child Care conducts community based active case finding. This activity is carried out by village health workers who screen the children for acute malnutrition in the community before the onset of complications using simple colored plastic strips known as Mid Upper Arm Circumference (MUAC) tapes. Village health workers also check for bilateral edema and refer children identified as undernourished to nearest health facility for further assessment and treatment as per the national protocol(5).

The treatment of moderate and severe acute malnutrition is supported by national integrated management of acute malnutrition (IMAM) guidelines and protocols adopted from WHO and UNICEF. The guidelines outline the admission, treatment and discharge criteria. The guidelines cover the four major components of community management of acute malnutrition namely community mobilization, supplementary feeding program, outpatient therapeutic care and in-patient therapeutic care. This aims at reducing mortality to less than 10%(4).

Children with MAM are referred to the supplementary feeding program where they receive nutrient-dense supplementary ration in order to meet their extra needs for weight and height gain and functional recovery(2). In addition breastfeeding promotion and support, education and nutrition counselling are provided to the caregivers of the child. The other activities include identification and prevention of the underlying causes of malnutrition, including nutrition insecurity(6).

Children with severe acute malnutrition (SAM) are screened for complications and those without complications are admitted to the outpatient therapeutic care (OTP), where they are provided with ready to use therapeutic foods (RUTF) while they are coming from home until adequate weight has been gained(7). The community-based approach involves timely detection of SAM in the community and provision of treatment for those without medical complications with ready-to-use therapeutic foods (RUTF) or other nutrient-dense foods at home and regular medical monitoring at a health facility(8).

Children with complicated severe acute malnutrition, who require more intensive support, are admitted to the Stabilisation Centre (SC) in the hospital and are treated for the complications.(8) As soon as the medical condition is stabilised and the oedema is reducing and the medical complication is resolving, the child is referred to outpatient care to continue the nutritional rehabilitation. It is expected that the child is in stabilisation for one to seven days(7).

The general principles of the WHO SAM treatment protocol are to treat and prevent hypoglycaemia, treat and prevent hypothermia, treat and prevent dehydration, correct electrolyte imbalance, treat and prevent infection and provide micronutrient supplementation(8).

The performance of quality IMAM program indicators (cure rate and defaulter rate) remains a challenge across the districts in the province. A preliminary review of district statistics of the first quarter of year 2017 showed that District A has highest acute malnutrition treatment defaulter rate at 67% against the target of less than 15% and low cure rate of 18, 6 % against the target of greater than 75 %.

Evidence of detailed review on IMAM program indicators has not been conducted from 2013 to 2018 in District A. Evaluating the IMAM indicator trends will help identify risk groups to inform decision making and planning regarding the health of the of the malnourished. We therefore carried out a secondary dataset analysis of the (IMAM) Dataset in District A to characterize the indicators of the acute malnutrition treatment

## Methods Study design

A descriptive cross sectional study was conducted on records from January 2013 to December 2018

### Study setting

The total population of this study area is estimated as 1,614,941 where 51% are females and 18% are children under 5 years of age. Approximately 2333 under-five SAM children were managed for acute malnutrition in this area for the year 2017. The study area in general 8 hospitals, 24 rural health centres. All health facilities in District A submitting IMAM data to HMIS were selected for record reviews.

### Study population

All malnourished patients admitted into IMAM at health facilities in District A, with a diagnosis of SAM from January 2013 – December 2018 as reported in the DHIS2 were used as the study population. Data from all the all health facilities within District A Districts were analysed.

### Data collection

District A IMAM data were retrieved from the DHIS2 to the excel sheet.

Sample size determination and sampling procedure

Since collated data from the DHIS and all records will be used for the study, no sample size calculation was conducted.

### Data analysis

Data were analyzed using Microsoft Office Excel 2007 to come up with graphs to demonstrate the various trends in IMAM from 2013 to 2018. R^2^ and p value were generated using statistics calculator.

### Permission to proceed

Permission to analyse data was sought from the National Nutrition Department, Health Studies Office and Midlands Provincial Medical Directorates’ office and District A Medical Officer.

### Ethical consideration

The data was handled with confidentiality and was only used for the purposes of this study. Stored data lacks patient names thus no personal information was accessed in this study and there was no need for signed consent forms in this study.

## Results

### Trends in total admissions into CMAM program in District A, 2013 to 2018

In 2013 371 cases / 100 000 were admitted into the CMAM program. In 2014 they slightly increased and declined slightly in 2015. This was followed by a sharp rise to a peak of 736 cases /100 000 in 2016. Cases admitted declined sharply in 2017 and slightly decreased again in 2018 reaching a lowest of 257 cases /100 000 in 2018.

### Trends in admissions by sex in District A District, 2013 to 2018

Figure 1 shows the trends in CMAM admissions by sex in District A District.Both sexes showed a general increase of admission cases in the CMAM program from 2013 to 2016, both reaching peaks in 2016. This was followed by steady decrease in admission of cases admitted in females (p =0.60 r square=0.07) to 104 cases/100 000 and unsteady decrease of male admitted cases to 153 cases/100 000 males (p=0.88 r square =0.005)

**Figure 1:**
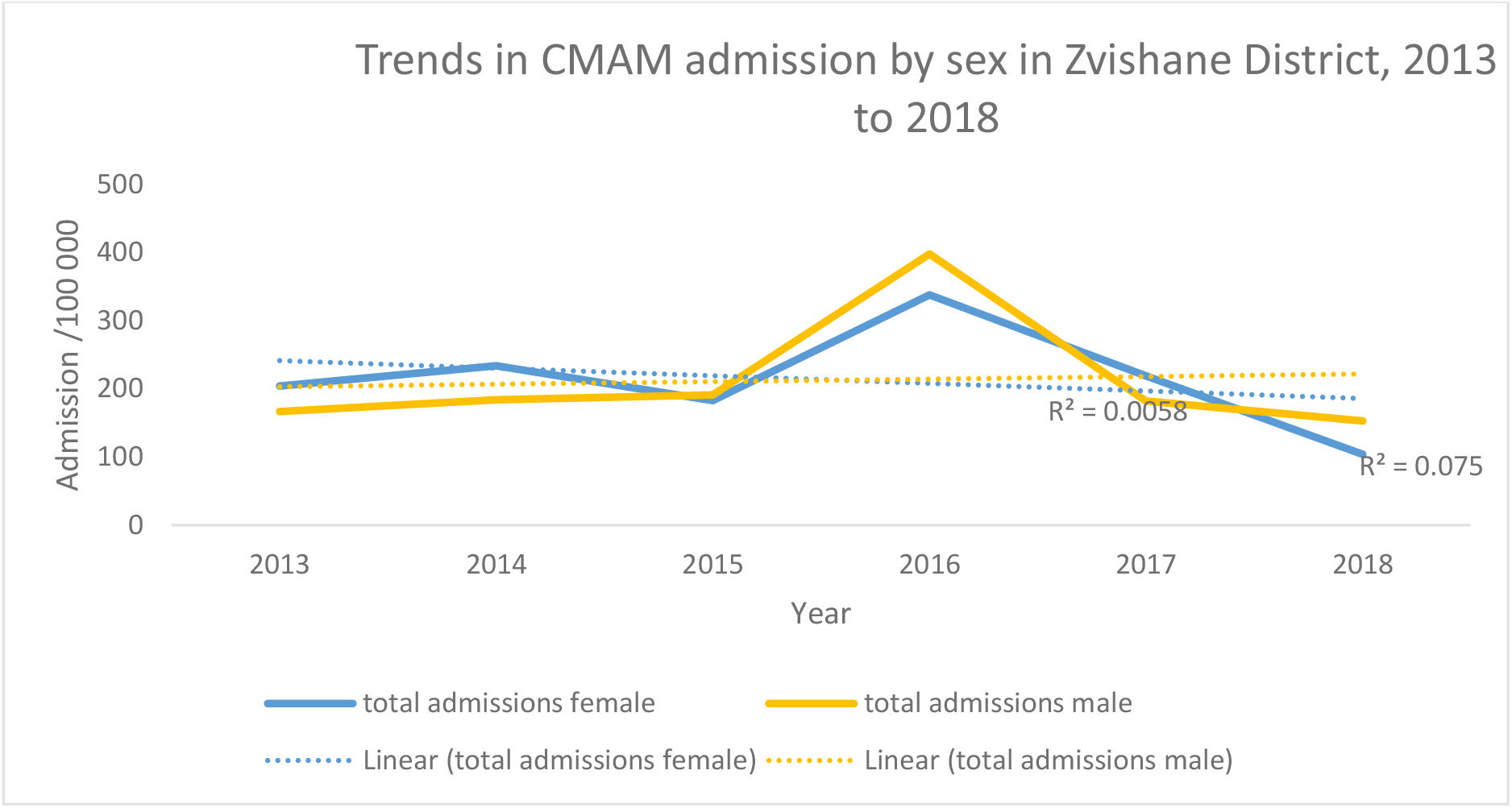
Trends in CMAM Admissions by sex District A District, 2013-2018

### Trends in CMAM Admissions by Age-groups, District A 2013-2018

Figure 2 illustrates the trends in CMAM Admissions by age-groups in District A 2013-2018. The 6-59 months age-group constitute the group with the highest cases of admission into the CMAM program. The cases in this particular age group showed an increase from 2013 to 2016 reaching a peak of 538/100 000 cases in 2016 and a general decrease from 2016 to 2018 reaching its lowest of 106cases/100 000. (p=0.8927, R^2^ =0.005)

**Figure 2:**
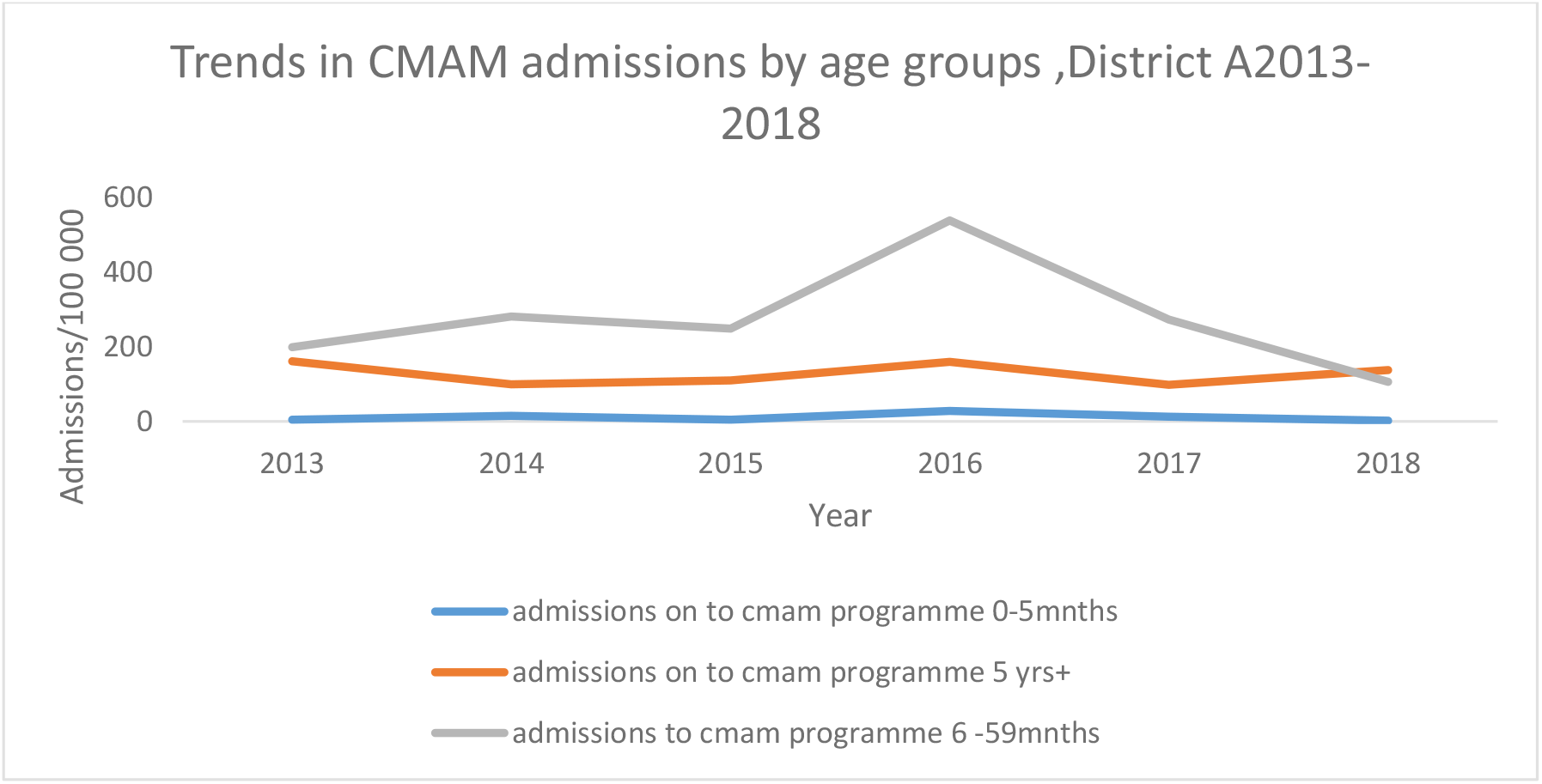
Trends in CMAM Admissions by Age-groups, District A2013-2018

The 0-5 months age-group produced a similar trend but at a reduced scale. This age group showed the least number of cases. (p=0.9369, R^2^=0.002)

The age group 5 years and above showed less number of admitted cases than the age group 6 to 59 months but on a higher scale than 0-5 month age group. The admissions in this age group were 199 cases / 100 000 in 2013. It slightly decreased to 99 cases/100 000 in 2014 followed by a slight increase from 2014 to 2016. Thereafter there was a slight decrease in the number of cases admitted in 2017 followed by a slight increase in 2018 reaching 138 cases/100 000. (p=0.8927, R^2^=0.003)

### Trends in CMAM admission by HIV positivity in District A, 2013-2018

In 2013 the HIV positivity was at 18%. The HIV positivity at admission decreased each year until 2016 which recorded the lowest positivity rate at 6%. A sharp increase was then noticed in 2017 followed by a slight increase in 2018 at 18%. (R^2^ =0.001 p =0.91)

### Trends in CMAM HIV positivity by admission and age in District A, 2013 to 2018

Displayed on the figure 3 are the trends in CMAM HIV positivity at admission by sex in District A

**Figure 3:**
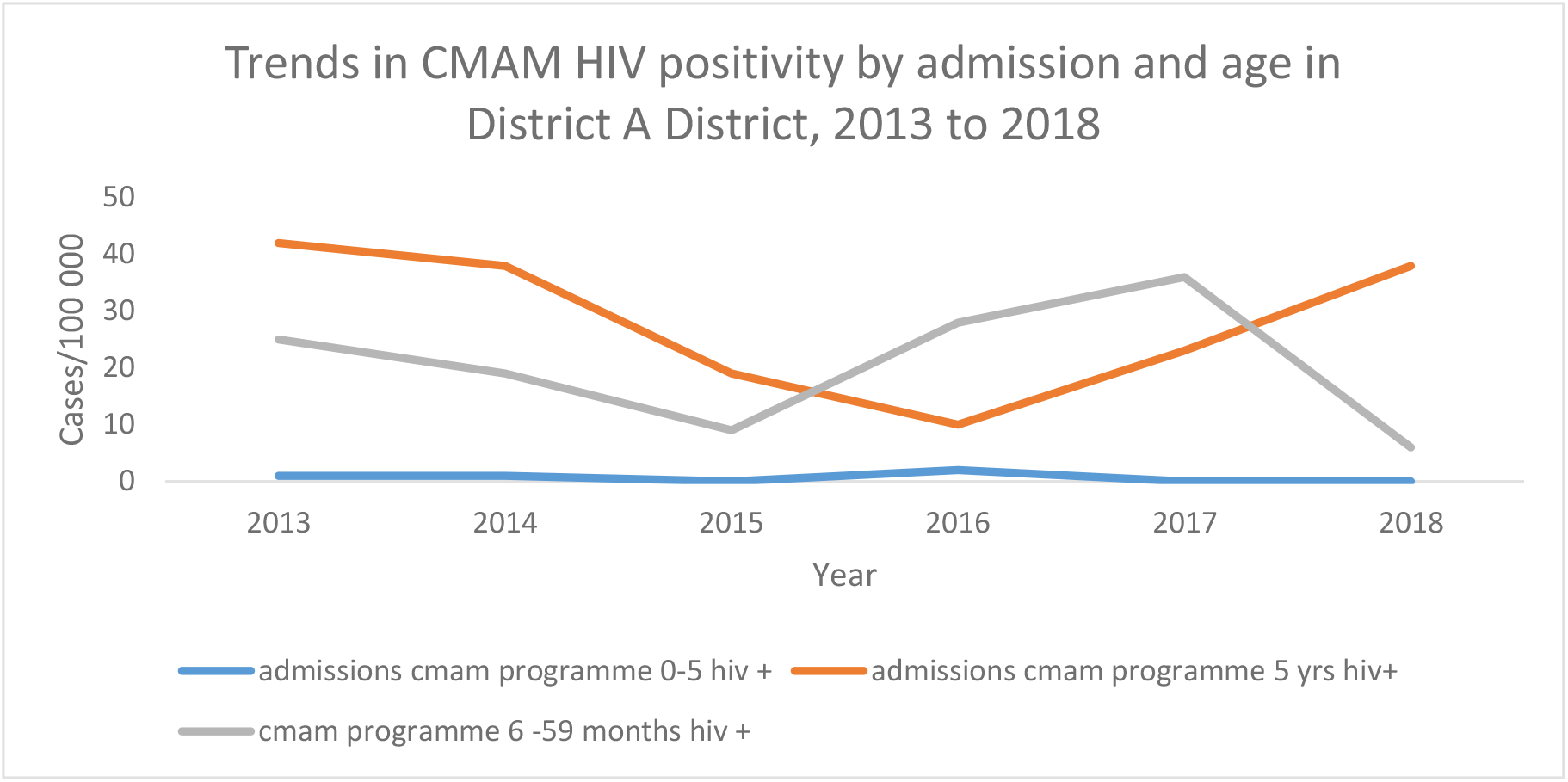
Trends in CMAM HIV positivity by admission and age in District A District, 2013 to 2018

The age group 0-5 months had the lowest HIV positivity over the period under review. In 2013 this particular age group had an HIV positivity rate of 1%. This remained constant up to the year 2015.A very slight increase was then noticed to reach 2% positivity which was the highest recorded in this age group. After this there was a decrease again to a 0% rate which remained constant up to 2018. (p=0.33, R^2^= 0.23)

The HIV positivity of the age group 5 years and above started the highest rate in 2013 at 42%. However it started to decline each year reaching its lowest rate of 10% in 2016. It then rose sharply from 2016 to 2018 to a peak of 38%. (p=0.56, R^2^ =0.09)

The age group 6 to59 month’s age group started with an HIV positivity of 25% in 2013. From there it started to decrease reaching its lowest HIV positivity in 2015 at 9%. It then sharply increased in 2016 to reach an HIV positivity of 28 % followed by a slight increase in 2017 to an HV positivity of 36%. A sharp decline was then experienced in 2018 its lowest rate of 6%. (p=0.83, R^2^ =0.013)

### Trends in total CMAM discharges in District A District, 2013 to 2018

In 2013 the least number of cases were discharged (193 cases/100 000) were discharged. There was a sharp rise in 2014 reaching a peak of 1420cases/100 000 followed by a sharp decline in 2015 reaching a lowest of 70 cases / 10000 in 2015. A slight increase in number of cases discharged was noticed in 2016 followed by a slight steady decline from 2016 to 2018. (p=0.58, R^2^=0.09)

### Trends in percent discharge from CMAM in District Afrom 2013 to 2018

In 2013 the lowest % discharge was recoded at 4%. This was followed by a sharp rise in the % discharge in 2014 and a sharp decline in 2015 which recorded the lowest % discharge in the period under review. The % discharges then experienced an upward trend from 2015 to 2018 reaching the highest % discharge in 2018 at 27%. (p=0.09, R^2^=0.53)

### Trends in CMAM discharges by sex in District A District, 2013 to 2018

The discharges in males (p=0.06 R^2^=0.06) and females (p=0.54 R^2^=0.06) showed a similar trend over the period, however female discharges were on a higher scale than males reaching a peak of 846 cases / 10000 in 2014 than males who reached a peak of 574cases/100000 in 2014.

### Trends in CMAM discharges by outcomes in District A2013 to 2018

Figure 4 displays the CMAM discharges by outcomes in District A District. In 2013 the death rate was lowest at 0.5% and there was a very slight increase in 2015 reaching rate of 4.2% which was the highest death rate for the period 2013 to 2018. Death rate slightly dropped from 2015 to 2016 and remained nearly constant from 2016 to 2018 at 0.9 %.(p=0.98 R^2^=0.00)

**Figure 4:**
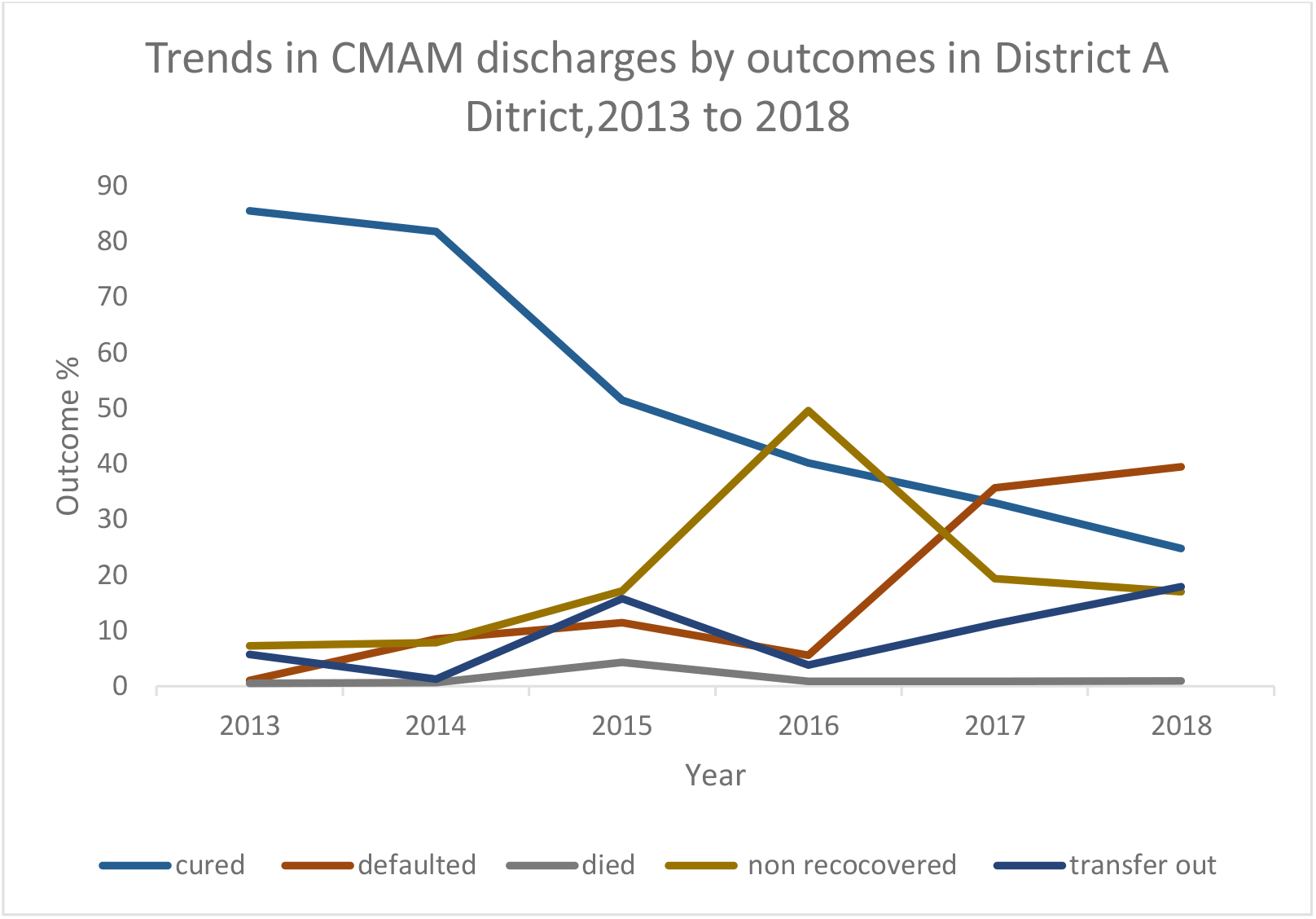
Trends in CMAM discharges by outcomes in District A2013 to 2018

The defaulter rate started at 1% in 2013. There was a gradual increase in the defaulter rate from 1% in 2013 to 11 % in 2015. After which the defaulter rate dropped slightly in 2016 followed by a sharp rise in 2017 reaching defaulter rate of the highest defaulter rate of 36 %. The rate increased again slightly reaching the highest defaulter rate of 39% in 2018. (p=0.03 R^2^=0.71)

In 2013 cure rate was the highest recorded outcome at 85%.The cure rate slightly dropped from 2013 to 2015 followed by a sharp decline in 2015. After which, it dropped gradually over the period from 2015 to 2018 recording the lowest cure rate of 25% in 2018. (p=0.005 R^2^=0.89)

In 2013 the rate of non-recovery was lowest at 7%. Thereafter there was a gradual increase from 2013 to 2015. This was followed by sharp increase in 2016 reaching a peak of 50%. From there it decreased sharply to a rate of 19% and slightly dropped again to a rate of 17% in 2018. (p=0.47 R^2^=0.13)

The rate of transfer out began at 6% in 2013. From there it slightly dropped to 1% in 2014, which was the lowest transfer out rate in the period. This was followed by a sharp increase to a rate of 15% and a sharp decline to a rate of 4% which was the lowest rate recorded in the period under review. A gradual increase then followed from 2016 to 2018 to reach a peak of 18% in 2028 which was the highest rate of transfer out recorded in the period under review. (p=0.2, R^2^=0.37)

### Trends in CMAM discharges by outcome and HIV positivity in District A District, 2013 to 2018

Figure 5 illustrates the trends CMAM discharges by outcome and HIV positivity in District Afrom 2013 to 2018.The highest outcome recorded among the HIV positive was death rate.

**Figure 5:**
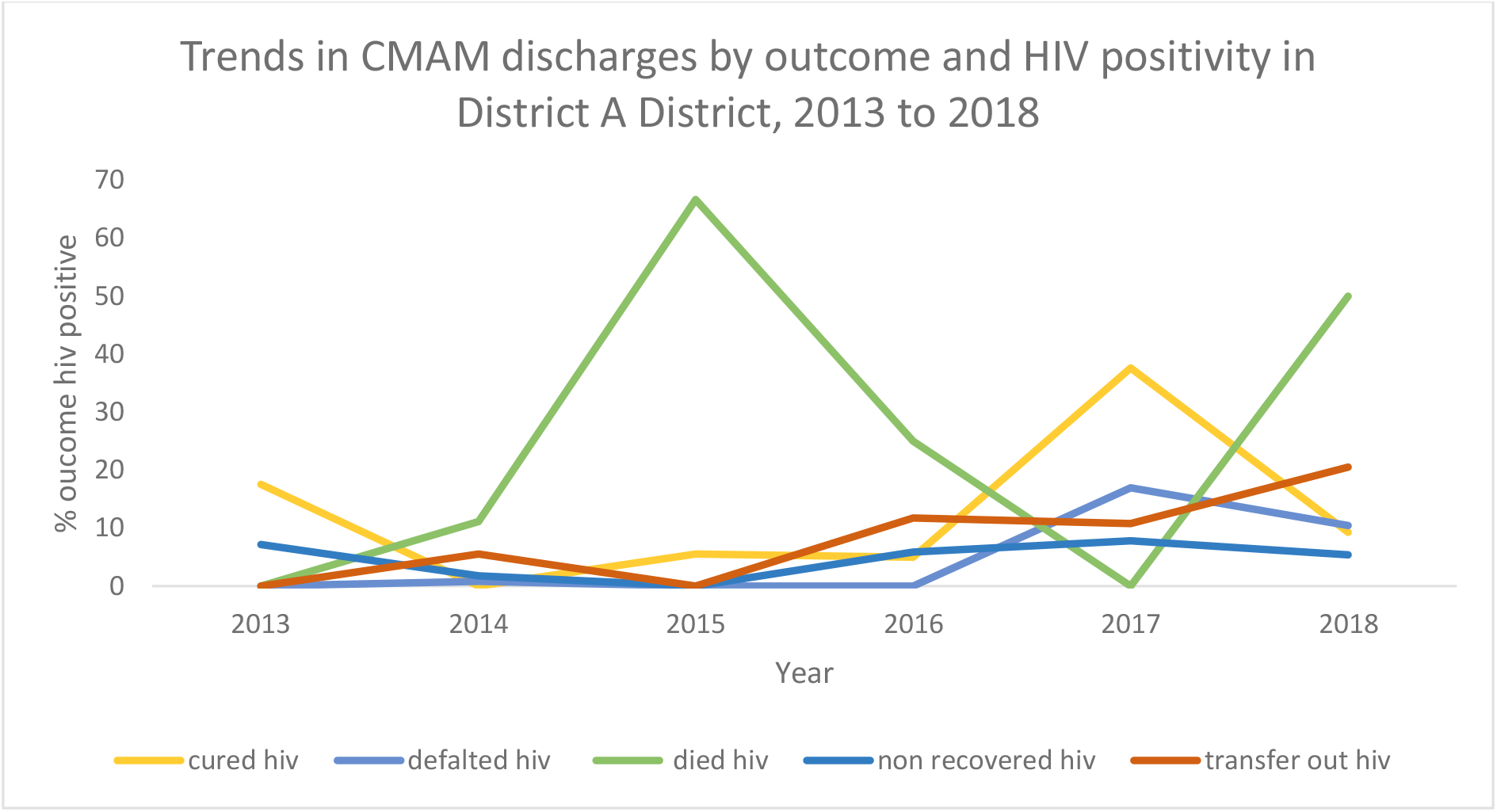
Trends in CMAM discharges by outcome and HIV positivity in District A District, 2013 to 2018

There was a steady increase of death rate in 2014 and a sharp increase in 2015, a general decrease from 2015 to 2017 and a sharp increase in 2018. (p=0.52, R^2^=0.11)

Transfer out started at 0%. However it showed an increase in the % transfer throughout the period under review with the highest % recorded in 2018 at 21%. (p=0.03 R^2^=0.72)

The rate of non-recovered started at 7% in 2013. It then showed a slight decline in 2014 and declined again in 2015 to reach the lowest non recovery rate of 0 %. This was followed by a gradual increase from 2016 to 2017. The rate decreased again slightly in 2018 to a rate of 5%. (p=0.65, R^2^=0.06)

The rate of defaulter in HIV positive started at 0% in 2013. The rate slightly increased to 0.8 % in 2014 and decreased to 0% in 2015 which remained constant up to 2016. A sharp increase was then noticed in 2017 to a rate of 17% which was the highest defaulter rate in the HIV positive. This was followed by a slight decline in 2018 to a rate of 10%. (p=0.14 R^2^=0.5)

The cure rate started at its highest in 2013 and dropped sharply to 0% in 2014. Thereafter a slight increase was noticed from 2015 to 2016. This was followed by a sharp increase in 2017 to its highest rate of 38%. The rate then sharply decreased to 9% in 2018. (p= 0.62 R^2^=0.07)

### Trends in CMAM therapeutic feeding, District A District, 2013 to 2018

Displayed on figure 6 is the therapeutic feeding program in District A District.The majority of CMAM therapeutic feeding involved RUTF from 2013-2018.In 2013 RUTF issued was 23645/100 000 units. The RUTF issued increased slightly in 2014 and the slightly reduced in 2015 followed by a sharp increase to a maximum of 40768 in 2017 then sharply decreased again in 2018 to 25189units/100 000. (p=0.40 R^2^=0.18).F75 formula was issued only in 2018 at 51 units. (p=0.17 R^2^=0.18). In 2013 1000 units of f100 were issued and sharply decreased to 451 units. No f100 formula was issued from 2015 to 2017.

**Figure 6:**
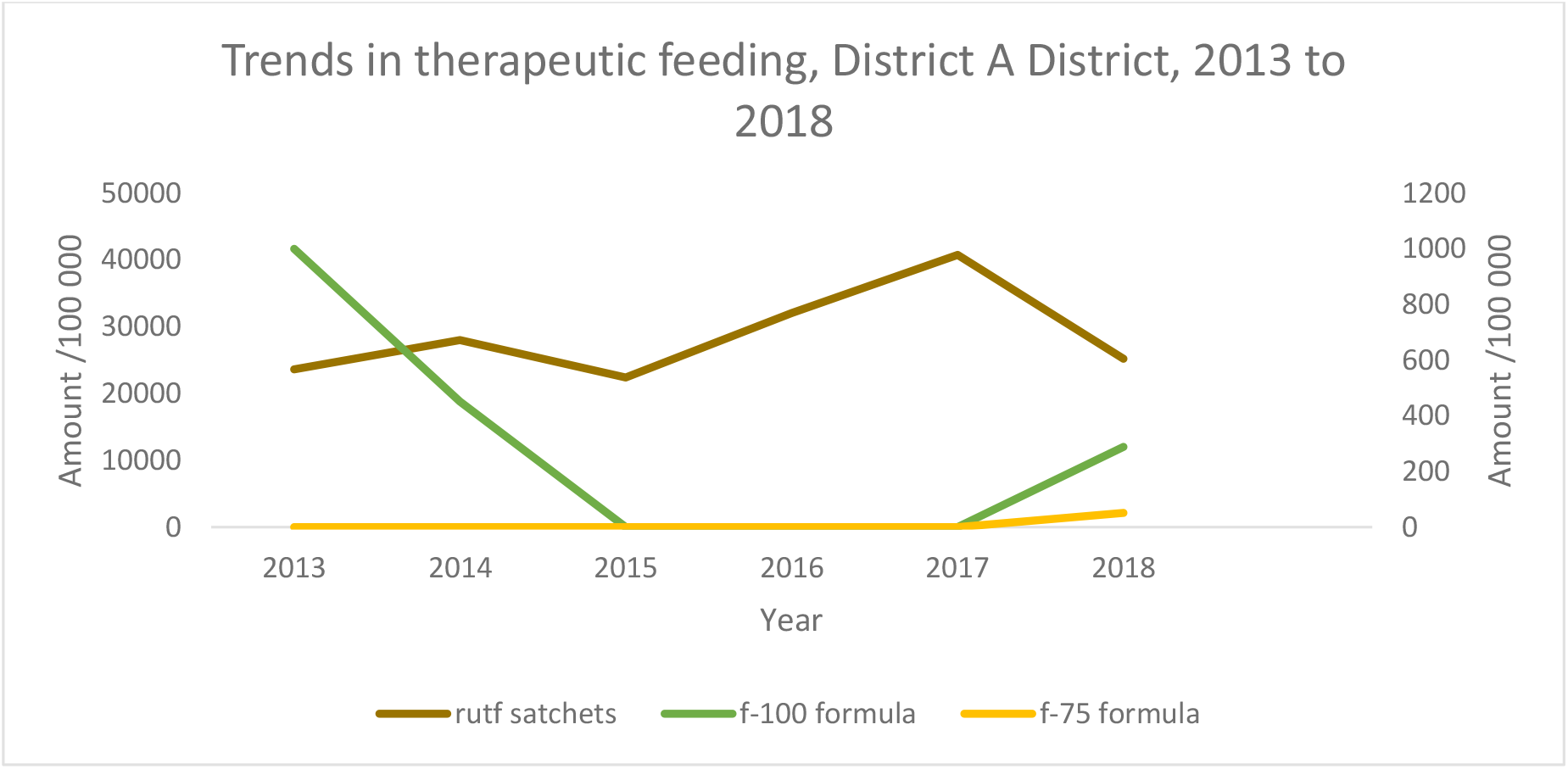
Trends in CMAM therapeutic feeding, District A District, 2013 to 2018

## Discussion

The aim of this study was to analyse trends of the management of acute malnutrition treatment and outcomes in District Afrom 2013 to 2018. This study showed that there was good quality data obtained from the district health information system which was available at 100% in the DHIS2.

The admission rates into the CMAM program were highest in 2016 across all age groups and in both males and females. This may have been due to high prevalence of food insecurity and inadequate diet. Intervention programs which were done in District Amay have resulted in the decrease in admission rate into the CMAM program. The district also received support for active screening which may have resulted in more children being enrolled into the program

.A study done in Nepal on food insecurity and vulnerability highlighted that factors like lack of access to care, lack of good sanitation and hygiene contributed to increased number of admission (9). The low admission in other years may be due to community mobilization and referral for early identification and treatment of SAM.

In this study malnutrition was prevalent in boys than in girls which was evidenced by the highest number of admitted cases (398/100 000) in males than (338/100 000) females. The higher prevalence of acute malnutrition among boys may be related to the higher growth rate in boys resulting in greater need for nutrients not supplied by diet. Wells et al in his study found that boys were more affected by environmental stress than girls which may have contributed to higher malnutrition in boys than in girls(10).

Contrary to our findings, a study done in India on gender disparities on the prevalence of undernutrition by Kshatriya et al, 2016 found that females were mostly affected by malnutrition than males although the reason why was not well explained(11).

In our study we found that the 6-59 months age-group constituted the group with the highest cases of admission into the CMAM program. This is similar to findings by Mittal et al in India who found higher prevalence of acute malnutrition among children (6-59 months) (12). This may be attributed to poor weaning and complementary feeding practices contributing to inadequate energy and protein intake.

According to WHO, 2018 few children receive nutritionally adequate and safe complementary foods. In many countries less than a fourth of infants 6–23 months of age meet the criteria of dietary diversity and feeding frequency that are appropriate for their age (13).

The age group 0 to 5 months had the least prevalence of malnutrition in our study. The possible reason is that majority of children in this age group are being breastfed with about 40% of infants being exclusively breastfed according to WHO, 2018.(14) For children above 5 years malnutrition is lower because these children can now start to consume food in the same patterns as adults.

As a standard indicator of CMAM programme performance, default rate ranks alongside mortality and cure rates as one of the principal measures of programme effectiveness(15). High default rates compromise the ability of a programme to respond to need and to achieve good coverage and hence a significant public health impact(16). Our study revealed that the performance of IMAM quality indicators remains a challenge despite investments. The district failed to meet the sphere standards for cure rate (>=75%) in the period from 2015 to 2018 and a defaulter rate (<15%) in the years 2017 and 2018.

In view of the fact that defaulting has continued to be problematic in District A District, there is need for, an in-depth investigation to better understand the reasons for and consequences of defaulting

A study done in Nigeria on the investigation of defaulting highlighted some reasons for high defaulter rate. The most common reason given by caregivers of defaulters who were interviewed were due to death and that mothers stopped coming because the children failed to eat the RUTF either because they were not able to swallow, did not like it or vomited. No money for transport and attitude of health workers were also cited as reasons which led to high defaulter rates(16).

A study by Al Amad et.al in the evaluation of outpatient therapeutic program for treatment of SAM in Yemen showed a high default rate among SAM children. Factors related were poor accessibility, poor satisfaction with staff and system, and treatment and acceptability of OTP. Having difficulty to attend OTP every week, unavailability of medication during follow-up visits, not liking to eat Plumpy’Nut were also cited as reasons for high default rate(17).

Bakwere etal in a study done in Ethiopia showed that the likelihood of recovery was 2.6 times higher for children with kwashiorkor than for those with marasmus(18). However our data do not provide specifications of whether on admission the child had oedema or not.

HIV and malnutrition often co-exist in children and malnutrition is a major problem for HIV infected children (19). Acute malnutrition was very common cause of admissions in all age groups. This could have been worsened by HIV. Acute malnutrition therefore remains a major global public health concern especially in the face of HIV.

The age group 0-5 months had the lowest HIV positivity over the period under review.This may be due to under reporting of HIV in the 0-5 months as DNA PCR needed for the less than 18 months is expensive. Another reason for the low prevalence of HIV in the very young is that most HIV –infected malnourished children die before their 2^nd^ birthday.

In our study the cure rate in HIV positive was low throughout the period recording less than 75% throughout the period. Similar to our findings Muluken et al in Ethiopia in a study on treatment outcome of severe acute malnutrition showed a significant association between low cure rate and the presence of HIV(20).

A systematic review and meta-analysis of HIV showed that more than 30% of severely malnourished children in sub-Saharan Africa admitted to inpatient nutrition rehabilitation units were HIV-infected(19). HIV-infected children with severe acute malnutrition were three times more likely to die compared with uninfected children. The infected group had persistent diarrhoea, pneumonia, extensive skin infections and oral thrush, which contributed to higher case fatalities and poorer response to management(19).

In our study, the highest outcome recorded among the HIV positive in CMAM program was death rate at 67% in 2015 followed by 50% in 2018. This showed that majority of those who are HIV positive and on treatment in the CMAM program died. The cure rate among the HIV positive in the CMAM program was also low and were below the sphere standard of (>=75%) throughout the period under review.

This shows that majority of HIV positive were not successfully cured in the CMAM program. A study in Malawi by Bawhere et al demonstrated that many HIV-infected children with severe acute malnutrition (SAM) can achieve an adequate Weight-For-Height (WFH) with appropriate therapeutic feeding, although recovery times are significantly longer and mortality is much higher compared to HIV-uninfected children(18). The possible reasons for slower weight gain include reduced intake due to poor appetite, nutrient malabsorption, increased incidence of infections that were unresponsive to the broad-spectrum antibiotics used, and increased nutrient requirements due to HIV.

Despite the possibility of reduced appetite especially at the beginning of treatment, HIV-positive children may need more RUTF than HIV-negative children to achieve similar growth rates and improvements in other nutritional indices. Increasing the amount of daily energy offered to HIV infected children may improve their weight gain and reduce the length of stay in the program. Further continued nutritional surveillance and supplementation after discharge may also help HIV-infected children to remain well-nourished.

## Conclusion

Male children were the majority of the patients admitted into the CMAM program. HIV positivity was highest in the 5 years and above age-group and less in children below 5 months. The treatment outcomes of this study showed that HIV negative had better treatment outcomes than HIV positive. The cure rate was lower than the sphere standard throughout the period under review among the HIV positive. High defaulter rates were noticed in 2017 and 2018.

## Recommendations

We recommended the district nutritionist to regularly monitor of the performance of IMAM program and to carry out support and supervision at clinics involved in CMAM program. There is also need to conduct a study on factors associated with high default rate in the CMAM program

## Data Availability

All data produced in the present work are contained in the manuscript

## Notes

### Competing Interest Statement

The authors have declared no competing interest.

### Funding Statement

This study did not receive any funding

